# ABO-RH blood group and risk of covid-19 in a moroccan population

**DOI:** 10.1101/2020.12.02.20242180

**Authors:** Mourad Belaouni, Elhoucine Malki, Rabii El Bahraouy, Bouchra El Maliki, Mohammed Er-Rami, Houcine Louzi, Khalid Lahmadi

## Abstract

**Introduction:** Given the rapid spread, significant morbidity and mortality associated with COVID-19, there has been scientific interest in obtaining data detailing the factors influencing the risk of COVID-19 infection. The aim of this study was to reveal a possible association between the ABO-RH system and the risk of COVID-19 in the Moroccan population.

**Materials and methods:** This is an analytical cross-sectional study. It was carried out on 1094 patients for the diagnosis of Covid-19 by Rt-PCR at the Moulay Ismaïl military hospital in the province of Meknes. All Rt-PCR negative individuals were used as a comparison group.

**Results:** Among the 1094 individuals who were diagnosed, RT-PCR for detection of SARS-CoV-2 was positive for 242 individuals. Comparison of the proportions of blood groups of the two groups showed that the proportion of blood group A in patients with COVID-19 was significantly higher than in people in the comparison group (P = 0.007), while the proportion of blood group O in patients with COVID-19 was significantly lower than in people in the control group (P = 0.017). Comparison of the Rh blood groups of the two groups did not find a significant association (P = 0.608).

**Conclusion:** As demonstrated by several previous studies, we concluded that blood group A was associated with a higher risk of acquiring COVID-19. Equally, the O blood group was associated with a lower risk of infection.

## Introduction

The 21st century has seen six global health emergencies declared by the World Health Organization (WHO), three of which were caused by viruses from coronavirus group. The most recent is a new Coronavirus. it was identified on January 7, 2020 and subsequently named SARS-CoV-2 (Severe Acute Respiratory Syndrome coronavirus 2). The disease was named COVID-19 (Coronavirus Disease 2019). March 11, 2020 the World Health Organization has increased the SARS-CoV-2 of an epidemic in a global pandemic [1].

Based on clinical observations, patient age, male gender and presence of certain underlying pathologies (primarily cardiovascular disease, chronic obstructive pulmonary disease, and diabetes) have been highlighted as risk factors predisposing to infection with SARS-CoV-2 and higher severity [2]. More recently, epidemiological and molecular studies seemed to report preliminary evidence on a correlation between the risk of contagion by SARS-CoV-2 and the erythrocyte phenotype determined by the ABO-RH system, the exact mechanisms of which are not yet well determined [3-4]. On the other hand, the association between blood groups and vulnerability or resistance to certain infectious pathologies has been demonstrated by older studies.

In this context, we conducted a study on the Association between ABO blood groups and COVID-19 infection in Moroccan population through patients tested for SARS-CoV-2 at the Moulay Ismail Military Hospital in Meknes.

## Materials and Methods

This is an analytical cross-sectional study involving male military people sharing the same workplaces, for the diagnosis of COVID-19 at the Moulay Ismail Military Hospital, in the province of Meknes, Morocco. The diagnosis was sought after the appearance of pulmonary symptoms in a few individuals.

On admission, all individuals underwent a complete physical examination and rapid serological test for the specific detection of IgM and IgG antibodies against the novel coronavirus. Sociodemographic, epidemiological and clinical data have been specified on information sheets. The detection of SARS-CoV-2 in nasopharyngeal and oropharyngeal samples was performed by real-time reverse transcription-polymerase chain reaction (RT-PCR) using the GeneFinder COVID-19 PLUS Real Amp Kit (OSANG Healthcare Co, Ltd., Korea) according to the manufacturer’s instructions, on the CFX96 real-time amplification system (Bio-Rad, Hercules, CA, USA). Testing for anti-Sars-Cov 2 antibodies was used to detect old infection (Rt-PCR negative), and in conjunction with epidemiological and clinical data for interpretation of questionable results. The ABO-RH blood typing was carried out on a sample taken from an EDTA tube using an agglutination technique using two complementary tests, Beth Vincent and Simonin. Our comparison group (COVID-19 negative) was made up of all individuals with RT-PCR and negative serology.

Statistical analysis was performed using Epi-Info (version 7.2 CDC Atlanta, USA). Proportions were compared using the chi-square or Fisher’s exact test. The means, meanwhile, were compared using Student’s test. A difference was statistically significant for P <0.05. An odds ratio (OR) with a 95% confidence interval was used as a measure of association.

## Results

This study involved 1094 individuals who were diagnosed with COVID-19. This is a relatively young male population. The average age was 36.32 years (standard deviation of 7.12 years, extremes ranging from 22 to 55 years). RT-PCR for detection of SARS-CoV-2 was positive for 242 individuals. 856 individuals with RT-PCR negative were used as a control group. The difference between the mean age of patients (35.13 years, SD = 7.189) and that of controls (36.66 years, SD = 1.070) was significant, which showed that the youngest in this community were at higher risk of SARS-CoV-2 infection (P = 0.003) (Table 1).

**Table 1:** Comparison between patients and controls with regards to ABO/Rh blood group

| | Control n=852 | COVID19 n=242 | $\chi^2$ | OR (IC à 95%) | $P$ |
| --- | --- | --- | --- | --- | --- |
| Age, mean (SD) | 36,66 (7,07) | 35,13 (7,189) | - | - | 0.003 |
| A (%) | 258 (30,28) | 96 (39,67) | 7,165 | 1,513 (1,125-2,035) | 0,007 |
| B (%) | 104 (12,21) | 32 (13,22) | 0,097 | 1,096 (0,716-1,676) | 0,754 |
| AB (%) | 48 (5,63) | 14 (5,79) | 0 | 1,028 (0,577-1,899) | 1 |
| O (%) | 442 (51,88) | 100 (41,32) | 5,643 | 0,697 (0,522-0,931) | 0,017 |
| RH+ (%) | 785 (92,14) | 226 (93,39) | 0,261 | 1,205 (0,685-2,121) | 0,608 |

The frequency distribution of blood groups A, B, AB and O in the patients was 39.67%, 13.22%, 5.79% and 41.32%, while in the controls it was 30.28%, 12.21%, 5.63%, and 51.88%, respectively. When blood groups Covid-19 patients were compared with the control group, it was found that The proportion of blood group A in patients with COVID-19 was significantly higher than that in control group (39.67% vs. 30.28%, P = 0.007), while the proportion of blood group O in Covid-19 patients was significantly lower than that of those in control group (41.32% vs. 51.88%, P = 0.017). These results revealed a significantly increased risk of COVID-19 for blood group A, with an odds ratio of 1.513 (95% CI 1.125-2.035) and a decreased risk of COVID-19 for blood group O, with an odds ratio of 0.697 (95% CI 0.522-0.931). Comparison of the Rh blood groups of the two groups did not find a significant association (P = 0.608) (Table 1).

Clinically, 17.36% (n = 42) of SARS-CoV-2 RT-PCR positive subjects were asymptomatic. All symptomatic patients presented with mild forms of COVID-19. The most frequent were anosmia and / or ageusia (n = 90), diarrhea (n = 80), cough (n = 62), sore throat (n = 60), breathing difficulties (n = 47), and asthenia (n = 44). The relationship between clinical expression of COVID-19 and the distribution of ABO blood groups was only significant for headaches. Covid-19 patients with group AB were more at risk of headache (p <0.001) (Table 2).

**Table 2:**
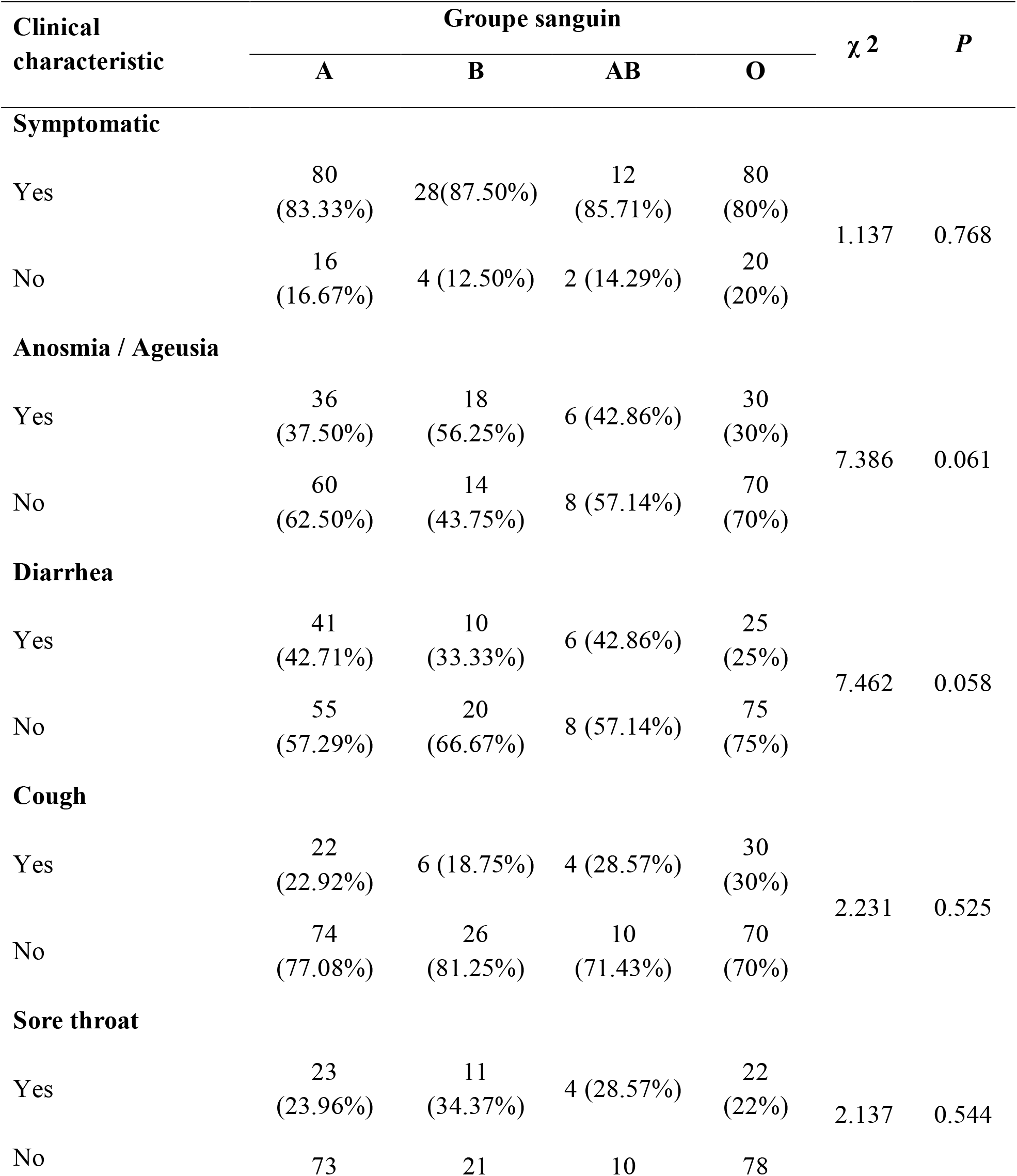
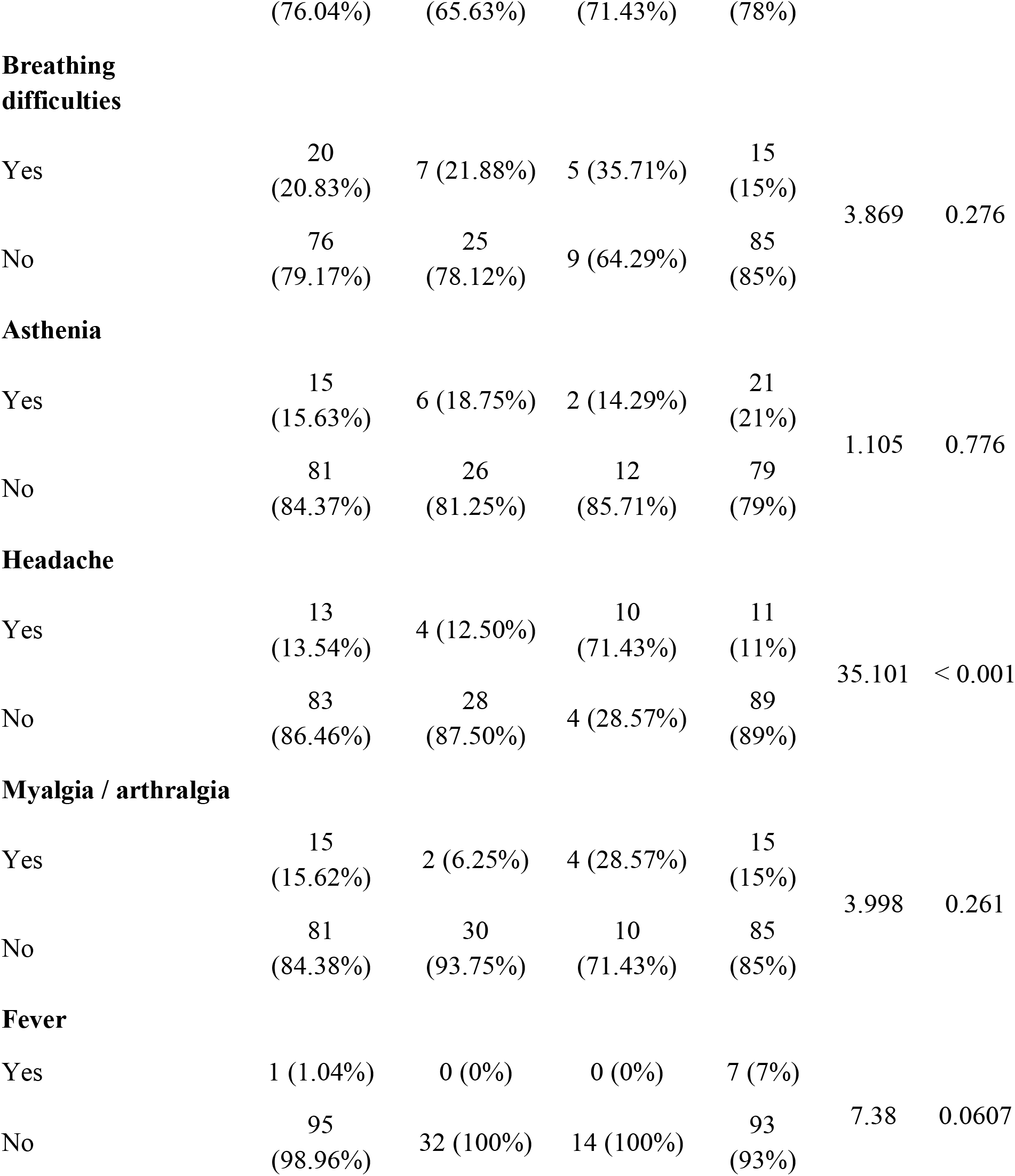
Relationship between distribution of ABO blood groups and clinical characteristic in patients with COVID-19

## Discussion

Similar to recently reported results, the objective of this work was to research an association between ABO-RH blood groups and risk of Covid19 in Moroccan population. And to our knowledge, our study is the first of its kind, in Morocco, to seek this association.

COVID-19 is the latest deadly zoonosis and, since its onset, it has affected millions of people around the world. So far, several risk factors, including advanced age, male gender, and presence of chronic underlying co-morbidities, have been established to be attributed to higher likelihood of being infected with SARS-Cov-2 [5-6]. Inconsistently with published data, we found that COVID-19 affects younger people more. This can be explained by the way of life and grouping which follow a hierarchical pattern. Indeed, the youngest are more in contact in workplaces and share guest rooms for four people or more, while the oldest (often older) share individual or two-person work spaces, and individual guest rooms.

Comparing the ABO blood groups of our COVID-19 patients with the control group, we found that there was a significantly increased risk of COVID-19 in patients with blood groups A and a significantly lower risk for patients with blood group O. This finding was consistent with the literature data [**7-**10]. Similar associations have been reported between ABO phenotypes and susceptibility to several bacterial and viral pathogens, where It can be vulnerability or resistance. It has been suggested that certain viruses perform their role by binding to ABO blood antigens. This is the example of Norwalk-like viruses and calciviruses which are spread by interaction with antigens of the ABO blood group system [**11-12**]. So for some strains of rotavirus, they find their way back through the cells of the gastrointestinal tract thanks to antigens associated with blood group A [13].

The association between the ABO blood group phenotype and the likelihood of being infected with SARS-CoV, the causative agent of severe acute respiratory syndrome (SARS), has been demonstrated. Yufeng Cheng et al. investigated the prevalence of SARS disease among hospital staff in Hong Kong who were exposed unprotected to infected patients, finding that people with blood group O had a lower likelihood of infection [14]. Jun Chen et al. proposed a mechanism by which the coronavirus spike protein (S), heavily N-glycosylated, bind with high affinity to the human angiotensin converting enzyme 2 (ACE2) [15]. As it has been shown that the ACE2 is a obligatory cell receptor, although other receptors may participate in the infection process [15-16]. Patrice Guillon et al. used in vitro cell binding tests and a mathematical model of cell viral transmission. They claimed that natural or monoclonal anti-A antibodies specifically inhibited the adhesion of the SARS-CoV (S) Spike protein to its ACE2 cell receptor [13]. Given the structural similarity between the receptor binding domains of SARS-CoV and SARS-CoV-2 [17] and also using the same receptor, ACE2, to enter target cells [17-18], the aforementioned mechanisms could be extended to SARS-CoV-2.

Our study did not find a clear association between the ABO blood group and clinical expression of COVID-19. Exception was made for headache where patients with AB blood type were more likely to be (P <0.001). Numerous studies published in this regard indicated that the ABO blood group does not influence the symptoms of COVID-19 [19-21].

This study had certain limitations which could lead to some bias in the results. First, the study population consisted exclusively of relatively young males. And second, the lack of other data (weight, underlying pathologies, smoking, etc.) which are predisposing factors to the risk of infection with SARS-Cov-2.

## Conclusions

As previous studies on COVID-19 and the present study have shown, the statistically significant association of the ABO blood group and susceptibility to COVID-19 is clear. Blood group A was associated with a higher risk of acquiring COVID-19 compared to non-A blood groups, while blood group O was associated with a lower risk of infection compared to non-O blood groups. The distribution of blood groups in the population can be useful to understand the kinetics of the epidemic at the local level and to establish a health policy aimed at reducing the viral spread. Further studies are needed to determine the exact mechanism by which the ABO blood group influences susceptibility to COVID-19, which could be useful in patient management and disease control.

## Data Availability

Data used to support the findings of this study are available from the corresponding author upon reasonable request.

